# Cardiometabolic Risk Factors for COVID-19 Susceptibility and Severity: A Mendelian Randomization Analysis

**DOI:** 10.1101/2020.08.26.20182709

**Authors:** Aaron Leong, Joanne Cole, Laura N. Brenner, James B. Meigs, Jose C. Florez, Josep M. Mercader

**Affiliations:** Department of Medicine, Harvard Medical School, Boston, Massachusetts, USA; Division of General Internal Medicine, Massachusetts General Hospital, Boston, Massachusetts, USA; Programs in Metabolism and Medical and Population Genetics, Broad Institute of MIT and Harvard, Cambridge, MA, USA; Diabetes Unit and Center for Genomic Medicine, Massachusetts General Hospital, Boston, MA, USA; Division of Endocrinology and Center for Basic and Translational Obesity Research, Boston Children’s Hospital, Boston, MA, USA; Division on Pulmonary and Critical Care, Massachusetts General Hospital, Boston MA

**Author notes:** These authors jointly directed this work (Aaron Leong, Josep M. Mercader). **Corresponding authors:** Aaron Leong, Massachusetts General Hospital, Division of General Internal Medicine, 100 Cambridge Street, 16^th^ floor, Boston, MA 02114, United States of America, Josep M. Mercader, Programs in Metabolism and Medical and Population Genetics, Broad Institute of Harvard and MIT, 75 Ames St, Cambridge, MA 02142, United States of America.

## Abstract

**Importance:** Early epidemiological studies report associations of diverse cardiometabolic conditions especially body mass index (BMI), with COVID-19 susceptibility and severity, but causality has not been established. Identifying causal risk factors is critical to inform preventive strategies aimed at modifying disease risk.

**Objective:** We sought to evaluate the causal associations of cardiometabolic conditions with COVID-19 susceptibility and severity.

**Design:** Two-sample Mendelian Randomization (MR) Study.

**Setting:** Population-based cohorts that contributed to the genome-wide association study (GWAS) meta-analysis by the COVID-19 Host Genetics Initiative.

**Participants:** Patients hospitalized with COVID-19 diagnosed by RNA PCR, serologic testing, or clinician diagnosis. Population controls defined as anyone who was not a case in the cohorts.

**Exposures:** Selected genetic variants associated with 17 cardiometabolic diseases, including diabetes, coronary artery disease, stroke, chronic kidney disease, and BMI, at *p*<5×10^-8^ from published largescale GWAS.

**Main outcomes:** We performed an inverse-variance weighted averages of variant-specific causal estimates for susceptibility, defined as people who tested positive for COVID-19 vs. population controls, and severity, defined as patients hospitalized with COVID-19 vs. population controls, and repeated the analysis for BMI using effect estimates from UKBB. To estimate direct and indirect causal effects of BMI through obesity-related cardiometabolic diseases, we performed pairwise multivariable MR. We used *p*<0.05/17 exposure/2 outcomes=0.0015 to declare statistical significance.

**Results:** Genetically increased BMI was causally associated with testing positive for COVID-19 [6,696 cases / 1,073,072 controls; *p*=6.7×10^-4^, odds ratio and 95% confidence interval 1.08 (1.03, 1.13) per kg/m^2^] and a higher risk of COVID-19 hospitalization [3,199 cases/897,488 controls; *p*=8.7×10^-4^, 1.12 (1.04, 1.21) per kg/m^2^]. In the multivariable MR, the direct effect of BMI was abolished upon conditioning on the effect on type 2 diabetes but persisted when conditioning on the effects on coronary artery disease, stroke, chronic kidney disease, and c-reactive protein. No other cardiometabolic exposures tested were associated with a higher risk of poorer COVID-19 outcomes.

**Conclusions and Relevance:** Genetic evidence supports BMI as a causal risk factor for COVID-19 susceptibility and severity. This relationship may be mediated via type 2 diabetes. Obesity may have amplified the disease burden of the COVID-19 pandemic either single-handedly or through its metabolic consequences.

**KEY POINTS:** *Question:* Is there a causal association between cardiometabolic conditions and COVID-19 susceptibility or severity?

*Findings:* Using two-sample Mendelian randomization of 17 cardiometabolic diseases and traits, only body mass index was found to be causally associated with testing positive for COVID-19 (6,696 cases/ 1,073,072 controls; *p*=6.7×10^-4^) and a higher risk of COVID-19 (3,199 cases/897,488 controls; *p*=8.7×10^-4^).

*Meaning:* Genetic evidence supports BMI as a causal risk factor for COVID-19 susceptibility and severity.

## INTRODUCTION

There is high heterogeneity in both susceptibility and severity of SARS-CoV2 infection with clinical severity^1,2^ ranging from asymptomatic carriers to life-threatening respiratory failure and death^3^. Epidemiological studies using both retrospective and prospective cohorts of different sizes and from multiple countries have reported evidence that underlying cardiometabolic conditions^4-29^ may be associated with an increased risk of severe COVID-19 illness (i.e., hospitalization, intubation, mechanical ventilation or death^30^). Coronary artery disease^4-6^, chronic kidney disease^7-12^, obesity^13-17^ and type 2 diabetes^8,18-21^ have a strong and consistent evidence for association with COVID-19 severity^30^. There is less compelling evidence for cerebrovascular disease^4,22-28^ (i.e., stroke) and hypertension^4,6,27-29^ leading to severe manifestations of COVID-19. Additional evidence suggests that these cardiometabolic traits may be associated with disease susceptibility^31^; however without universal testing, this correlation is difficult to prove.

While early reports are crucial to inform clinical decision making and public health policy during a pandemic of a new pathogen, correlative observational data can be plagued by residual confounding. Thus, there remain inherent challenges in drawing causal inferences from these epidemiologic studies. Mendelian Randomization (MR) is an analytic approach that uses human genetic variation known to influence modifiable exposures to examine their causal effect on disease^32^. MR is especially useful for disentangling causal pathways of phenotypically clustered risk factors that are difficult to randomize or prone to measurement error. By identifying causal relationships between cardiometabolic risk factors and COVID-19 susceptibility or severity, we may be able to mitigate their impact on disease risk and avoid spurious conclusions that lead to misinformation or incite unnecessary anxiety.

We hypothesize that only some cardiometabolic conditions have a causal relationship with COVID-19 illness or its disease course. Thus, we sought to evaluate the causal associations of 17 cardiometabolic exposures with COVID-19 susceptibility and severity using two-sample MR analyses. Causal effects were estimated from genome-wide association studies (GWAS) summary statistics of these cardiometabolic diseases and related traits and COVID-19-related outcomes from the COVID-19 host genetics initiative (https://www.covid19hg.org/)^33^.

## METHODS

### Candidate instrument selection for cardiometabolic diseases and traits

We extracted association summary statistics from published large-scale GWAS meta-analysis to generate sets of genetic instruments for 17 cardiometabolic diseases and traits, type 1 diabetes^34^, type 2 diabetes^35^, hemoglobin A1c^36^, fasting glucose adjusted for body mass index (BMI)^36^, fasting insulin adjusted for BMI^36^, BMI^37^, waist-hip ratio^38^, low-density lipoprotein cholesterol^39^, high-density lipoprotein cholesterol^39^, triglycerides^39^, systolic blood pressure^40^, diastolic blood pressure^40^, creatinine-based estimated glomerular filtration rate (eGFR)^41^, chronic kidney disease^41^ coronary artery disease^42^, any stroke^43^, and c-reactive protein^44^ (CRP), a non-specific biomarker of inflammation that can be elevated in people with high cardiometabolic risk. We used genetic variants associated with these exposures at genome-wide significance (*p*<5×10^-8^) and excluded those that were not represented in the COVID-19 outcome GWAS datasets. Using the LD_clumping function, we pruned the list of candidate instruments for linkage disequilibrium (LD; R^2^>0.01) and discarded variants that were within 1-Mb distance from other candidate instruments with a stronger association. Aanalyses were performed using the R package “twosampleMR” v.4.0^45,46^.

### COVID-19 susceptibility and severity from the COVID-19 Host Genetics GWAS meta-analysis

The COVID-19 Host Genetics Initiative is an international genetics collaboration that aims to uncover the genetic determinants of outcomes related to COVID-19 susceptibility and severity^33^. To accomplish this, investigators from around the world assembled individual-level clinical and genetic data and performed individual GWAS. Summary statistics were shared via a cloud-based computing platform, and centralized meta-analysis was performed. Single-variant association testing were adjusted for age, age^2^, sex, age*sex, genetic ancestry principal components and other study-specific covariates. An allele frequency filter of 0.0001 and an INFO filter of 0.6 was applied to each study prior to meta-analysis with inverse-variance weighting. Summary statistics from the third round of GWAS meta-analysis, shared publicly on July 2, 2020, were used to test the 17 sets of genetic instruments against COVID-19 outcomes assembled by the COVID-19 Host Genetics Initiative.

For our two primary analyses, we selected the COVID-19 outcomes with the largest number of cases. For Susceptibility: 1) COVID-19 by RNA PCR, serologic testing, or clinician diagnosis by chart review or ICD-coding (6,696) vs. population controls (N=1,073,072) and for Severity: 2) hospitalization of patients with COVID-19 by RNA PCR, serologic testing, or physician diagnosis (N=3,199) vs. population controls defined as any person who was not a case (i.e., people who tested negative, were never tested, or had an unknown testing status; N=897,488),As controls were not selected based on testing results, specific characteristics, or testing status, they were likely to be representative of the general population.

To determine whether statistically significant results from the primary analyses were consistent across different definitions for COVID-19 susceptibility, severity, and control groups, we performed secondary MR analyses of the four remaining available outcomes. For Susceptibility: 1) COVID-19 positive by RNA PCR, serologic testing, or clinician diagnosis (N=3,523) vs. COVID-19 negative by RNA PCR, serologic testing, or self-report (N=36,634); 2) predicted COVID-19 based on symptoms or COVID-19 positive by self-report (N=1,865) vs. no predicted COVID-19 based on symptoms or no COVID-19 by self-report (N=29,174) using a model developed by Menni et al. 2020^47^, and for Severity: 3) critical respiratory illness, defined by death, intubation, Continuous Positive Airway Pressure (CPAP), Bilevel Positive Airway Pressure (BiPAP), Continued external Negative Pressure (CNP), or very high flow positive end expiratory pressure oxygen in patients with COVID-19 by RNA PCR or serologic testing (N=536) vs. population controls (N=329,391) and 4) hospitalization (N=928) vs. no hospitalization within 21 days of testing positive for COVID-19 (N=2,028).

### Mendelian Randomization analysis of COVID-19 susceptibility and severity

To estimate the causal association of each exposure with each outcome, we performed two-sample MR analyses using the random-effects inverse-variance weighted (IVW) method, whereby genetic variant-outcome coefficients were modeled as a function of genetic variant-exposure coefficients weighted by the inverse of the squared genetic variant-outcome standard errors^48^. The use of random effects provides a concise estimation and considers potential heterogeneity among estimates from individual variants^49^. We used *p*<0.05 / 17 exposures / 2 outcomes = 0.0015 to declare statistical significance with the understanding that this threshold may be conservative as exposures are clinically correlated. We reported causal effect estimates as odds ratios for the outcome per log-odds of binary exposures or unit change of continuous exposures. For BMI, we repeated the analysis using untransformed variables from UK Biobank (http://www.nealelab.is/uk-biobank) to report causal effect estimates per unit change of raw BMI.

### Accounting for pleiotropy

An assumption of MR is that instruments do not influence the outcome independently of the risk factor of interest (i.e. non-mediated pleiotropy). We tested this assumption in a series of sensitivity analyses. We used the Weighted Median Estimator (WME)^50^ which requires ≥ 50% of the contribution to the causal estimate to be from valid instruments; if so, its causal estimate is stable. We then used the MR-Egger regression^51^ whereby a linear regression of variant-outcome on variant-exposure coefficients was performed without constraining the intercept to the origin. The slope of the regression line provides the corrected causal estimates even when none of the instruments are valid^51^. Next, we used the mode-based estimate which is consistent when the largest number of similar single-variant MR estimates are derived from valid instruments even when the majority are invalid^52^. If all MR models produce similar causal estimates despite making different assumptions on the validity of instruments, we would be more confident of the robustness of our results^53^. In other sensitivity analysis, we applied MR pleiotropy residual sum and outlier (MR-PRESSO)^54^ and leave-one-out analysis to determine whether outliers may be biasing the overall causal estimate. To estimate direct and indirect causal effects of BMI via obesity-related cardiometabolic diseases, CAD, stroke, CKD, type 2 diabetes, and CRP, we performed pairwise multivariable MR wherein we conditioned upon the effects of these exposures with BMI to simultaneously estimate their independent causal effects.

## RESULTS

### Selection of genetic instruments for exposures

We obtained genetic instruments for the 17 exposures for MR analyses after excluding variants that were in LD (r^2^>0.01) and in close proximity (1 Mb) to other candidate instruments with stronger *P*-values. Genetic instruments explained between 0.2 to 5.3% of the variance or liability of each exposure (Table 1). Contributing studies included in these exposure GWAS meta-analyses were predominantly of European ancestry.

**Table 1.** Candidate genetic instruments of cardiometabolic diseases and traits.

| Exposure | PMID | Sample size, N | Ancestry | Candidate genetic instruments, N | Genetic instruments used in analysis, N | Estimated variance explained (%) |
| --- | --- | --- | --- | --- | --- | --- |
| Type 1 diabetes | 25751624 | 6,808 cases/ 12,835 controls | European | 75 | 50 | 3.2 |
| Type 2 diabetes | 30297969 | 898,130 (9% cases) | European | 243 | 226 | 3.1 |
| A1C | Chen J, BioRxiv 2020 | Up to 281,416 | 70% European | 216 | 105 | 2.2 |
| FG | Chen J, BioRxiv 2020 | Up to 281,416 | 70% European | 179 | 91 | 1.6 |
| FI adjusted for BMI | Chen J, BioRxiv 2020 | Up to 281,416 | 70% European | 96 | 61 | 1.0 |
| BMI | 25673413 | Up to 339,224 | Mostly European | 75 | 72 | 1.7 |
| WHR adjusted for BMI | 25673412 | Up to 224,459 | Mostly European | 53 | 43 | 0.8 |
| CRP | 31900758 | Up to 418,642 | European | 439 | 437 | 5.3 |
| LDL | 24097068 | Up to 188,577 | European | 65 | 63 | 1.9 |
| HDL | 24097068 | Up to 188,577 | European | 54 | 53 | 1.8 |
| Triglycerides | 24097068 | Up to 188,577 | European | 39 | 38 | 1.3 |
| Systolic blood pressure | 30224653 | >1,000,000 | European | 185 | 181 | 1.5 |
| Diastolic blood pressure | 30224653 | >1,000,000 | European | 190 | 183 | 1.5 |
| Creatinine-based eGFR | 31152163 | >1,000,000 | Mostly European | 547 | 280 | 3.3 |
| Chronic kidney disease | 31152163 | 64,164 cases/561,055 controls | Mostly European | 23 | 21 | 0.6 |
| CAD | 28714975 | 10,801 cases/137,914 controls | Mostly European | 50 | 50 | 0.9 |
| Any stroke | 29531354 | 67,162 cases and 454,450 controls) | Mostly European and East Asian | 23 | 16 | 0.22 |

Where available, we used European-specific effect estimates in the MR analysis. Sample sizes were the maximum number indicated in the published manuscript. Estimated variance explained by genetic instruments was a sum of estimated variance explained by each variant calculated from reported *P*-values, sample sizes, and proportion of cases and controls using the TwoSampleMR R functions `get_r_from_lor()` and `get_r_from_pn()`.

### Causal effect of each cardiometabolic exposure on COVID-19 susceptibility and severity

Of the 17 cardiometabolic exposures, only BMI was found to be causally associated with COVID-19 susceptibility and severity even after accounting for multiple testing (Figure 1). Genetically increased BMI was associated with a higher risk of testing positive for COVID-19 (*p*=6.7×10^-4^) and a higher risk of COVID-19 hospitalization (*p*=8.7×10^-4^) compared to population controls using random effects IVW (Figure 1). For both outcomes, we identified no heterogeneity of effects (*p*=0.52; *p*=0.49,) or outlying genetic variants by the leave-one-out analysis or MR-PRESSO. To obtain interpretable effect estimates, we repeated the analysis using beta estimates of raw BMI from UK Biobank^55^ and found consistent results: odds ratio 1.08 per kg/m^2^ increase in BMI (95% CI 1.03, 1.13, *p*=1.3×10^-3^ for testing positive with COVID-19; odds ratio 1.12 per kg/m^2^ increase in BMI (95% CI 1.04, 1.21, *p*=1.7×10^-3^) for COVID-19 hospitalization. Point estimates from the MR-Egger, WME, and weighted MODE, were in the same direction as IVW (Figure 2 and Supplemental Figures 1-6). The MR-Egger intercept *p* was 0.49 and 0.24 for susceptibility and severity, respectively, indicating the absence of directional pleiotropy. The MR results of the remaining four COVID-19 susceptibility and severity outcomes had *p*>0.001 (Supplemental Table 1).

**Figure 1.**
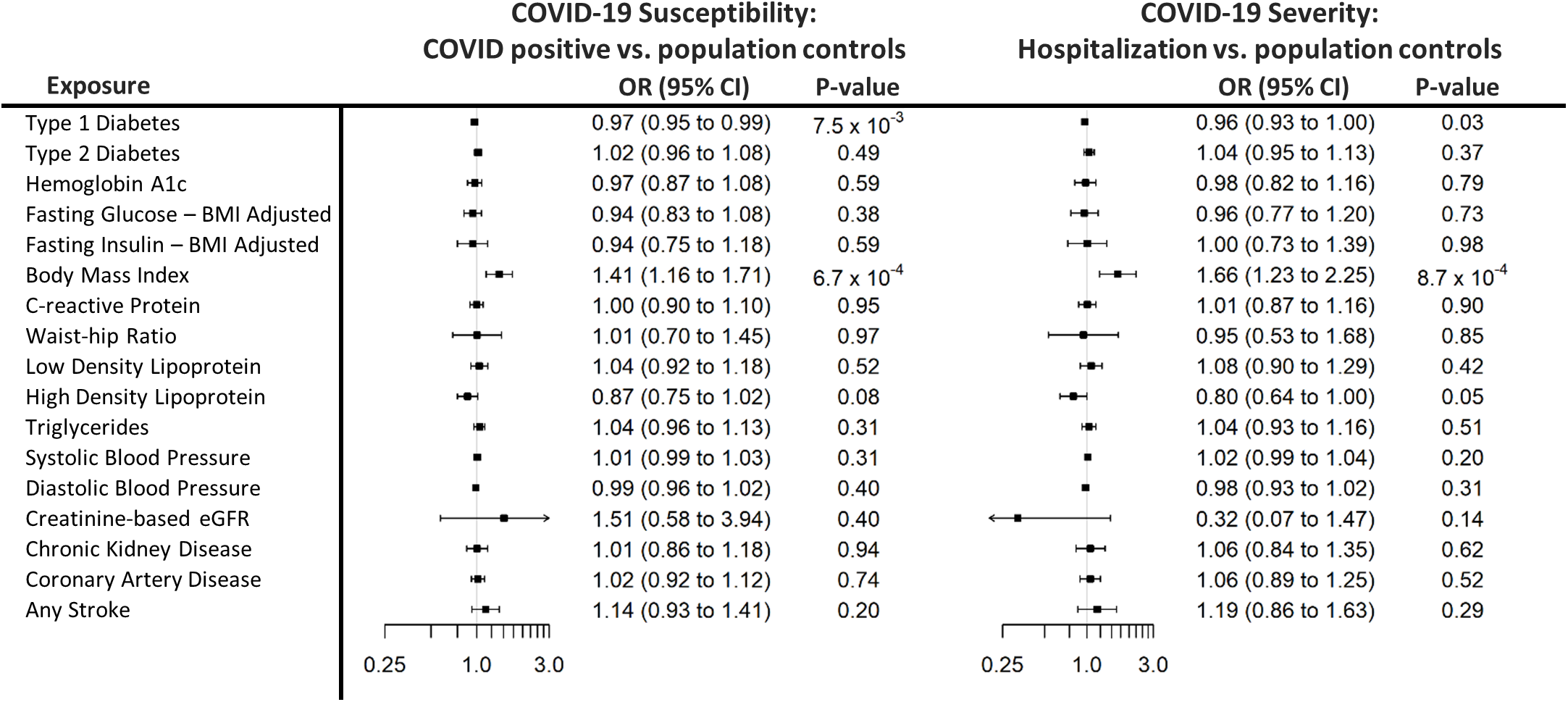
Forest plot causal effect estimates and 95% confidence interval for each exposure and the two main outcomes analyzed. Causal estimates are reported as odds ratios per unit of the exposure: hemoglobin A1c, A1C: %-unit; fasting glucose, FG: mg/dL; fasting insulin, FI: natural log; body mass index, BMI: inverse normally transformed residuals; waist-hip-ratio, WHR: inverse normally transformed residuals; c-reactive protein, CRP: rank-based inverse-normal transformed; low-density lipoprotein, LDL: standardized; high-density lipoprotein, HDL: standardized; triglycerides, TG: standardized; systolic and diastolic blood pressure: mmHg: eGFR ml min-1 per 1.73 m^2^; type 1 diabetes, type 2 diabetes, coronary artery disease, chronic kidney disease, any stroke: log-odds.

**Figure 2.**
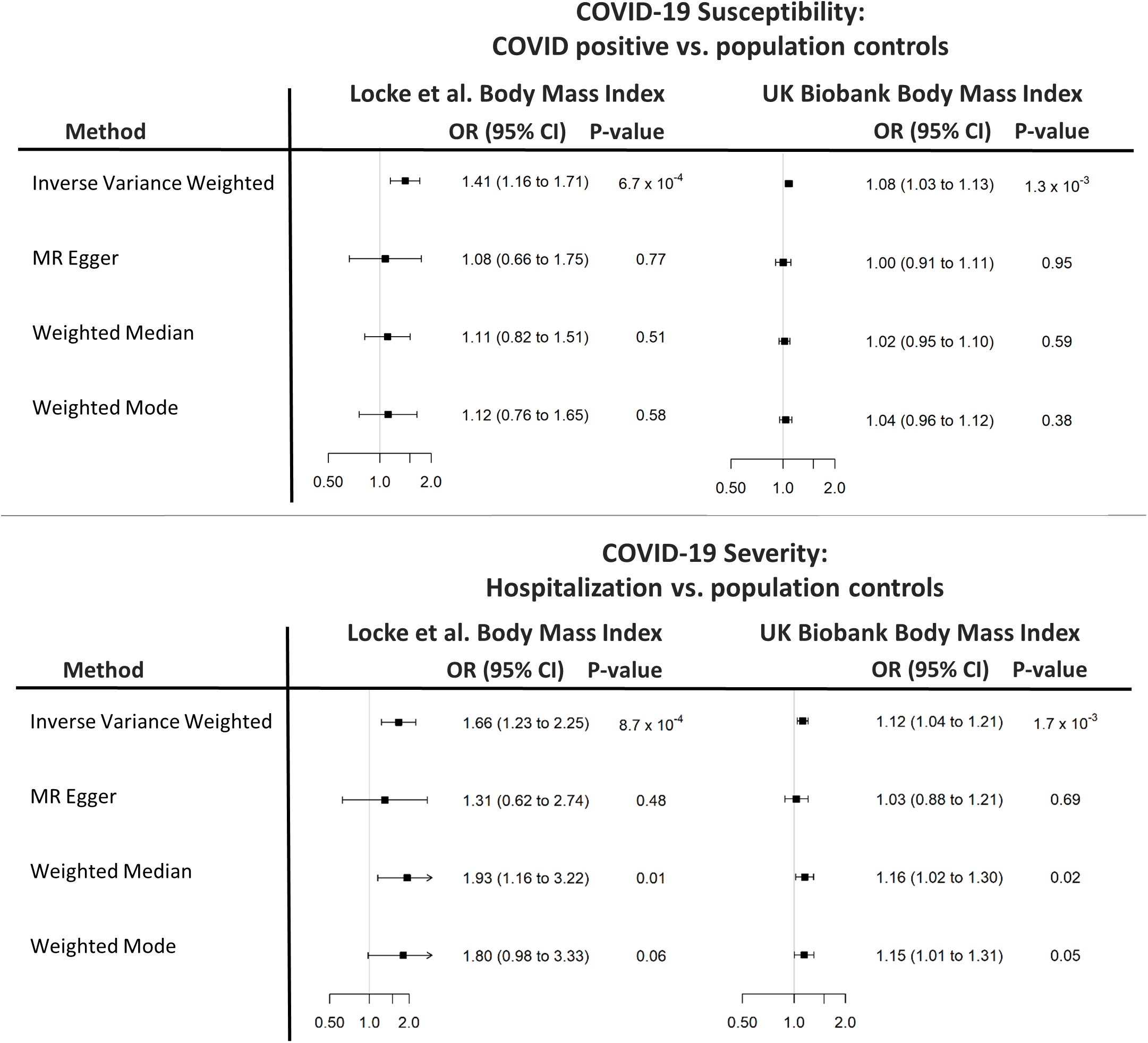
Sensitivity analyses using other MR methods and results using UK Biobank effect estimates. Causal estimates were reported as odds ratios (OR) per unit increase in body mass index (BMI). Locke et al.: inverse normally transformed residuals; UK Biobank: kg/m^2^

To determine whether the causal effect of BMI was mediated through obesity-related cardiometabolic diseases, we performed pairwise multivariable MR of BMI with each of the cardiometabolic diseases, type 2 diabetes, chronic kidney disease, coronary artery disease, any stroke, and CRP. The direct effects of BMI on the two COVID-19 outcomes were abolished upon conditioning on the genetic effects of type 2 diabetes (*p*>0.05). Adjusting for the genetic effects on the other diseases did not attenuate the direct effect of BMI (*p*<0.05, Supplemental Table 2)

While none of the other cardiometabolic exposures was found to increase COVID-19 susceptibility or severity, we found a borderline association between having a higher genetic predisposition to T1D with a *lower* risk of testing positive for COVID-19 and hospitalization vs. population controls, though the associations were not statistically significant after accounting for multiple testing (Supplemental Table 3).

## DISCUSSION

Cardiometabolic diseases have been identified to be risk factors for COVID-19 illness^30^. Since risk factors may be only correlated, and not causally related, with outcomes of interest, it is paramount to assess causality to inform preventive strategies. Using two-sample MR, we found that genetically increased BMI was the only risk factor for COVID-19 susceptibility and severity among the 17 cardiometabolic diseases and traits tested, whereby the odds of testing positive for COVID-19 was 8% higher per kg/m^2^ increased in BMI and the odds of hospitalization with COVID-19 was 12% higher per kg/m^2^ increase in BMI than the general population. Our MR findings support the multiple epidemiologic studies that have reported a strong and robust association between obesity and COVID-19 illness^13-17^. Adjusting for the genetic effect of type 2 diabetes obliterated the direct causal effect of BMI, suggesting that type 2 diabetes may be a mediator in the causal association of BMI and COVID-19 illness. By understanding causality, we can aim to modify causal exposures for the purpose of mitigating disease risk.

Apart from BMI, the other cardiometabolic exposures tested are unlikely to play a key causal role in contracting COVID-19 or worsening the illness. Observational correlations of cardiometabolic conditions with COVID-19 outcomes may be partly due to clinical clustering with obesity. It is noteworthy that correlational risk factors can still have clinical utility in identifying at-risk patients even if causality is refuted. However, if preventive efforts only target correlated, but not causal, risk factors, disease risk may not be reduced. We found a negative trend between a higher genetic predisposition for type 1 diabetes and a lower risk of hospitalization and testing positive for COVID-19. A negative association with COVID-19 outcomes could be observed if people with underlying medical conditions were more likely than the general population to undergo testing for COVID-19 and receive a negative test result, made concerted efforts at reducing their risk of viral exposure in response to public health messaging, or were encouraged by health professionals to recuperate at home and not come to the hospital when ill with mild viral symptoms. We could speculate that the autoimmune nature of type 1 diabetes provides protection compared to the general population. Additionally, it is noteworthy that people with T1D, unlike T2D, do not generally have higher BMI than the general population. More investigation is needed to further understand the cause of this negative association.

Our study had limitations. The variances explained in the exposures by genetic instruments were modest, though well within the ranges that were typical for complex traits. The use of weak genetic instruments could have limited our ability to detect subtle causal associations and does not exclude the possibility of modest effects. It is also possible that, with larger sample sizes, the association of other cardiometabolic exposures with COVID-19 outcomes may become significantly significant and confidence intervals would narrow around true estimates. Additionally, our analysis did not factor non-linear exposure-outcome relationships. The causal estimates by MR-Egger were not as compelling suggesting that horizontal pleiotropy or other confounding factors could have biased estimates. Yet, MR-Egger is a less efficient estimator than the other methods^50^ and is generally considered as only one of several sensitivity analyses used to evaluate the plausibility of findings. In our primary analyses we chose to use controls that were broadly defined as not being a case. Without universal testing, the control group, albeit representative of the general population, could have been contaminated with people who had contracted COVID-19, particularly those with only mild or no viral symptoms (asymptomatic), which would have biases estimates towards the null. Nevertheless, our results were consistent when using controls that were narrowly defined as people who tested negative for COVID-19.

Obesity contributes to higher levels of circulating proinflammatory adipokines and cytokines^56-61^ which may intensify virally induced inflammation,^62-69^ and could contribute to acute respiratory distress syndrome, the main cause of mortality from COVID-19^70,71^. We did not include critical respiratory illness in the primary analysis because the sample size of cases was small. When larger samples become available, future MR analyses can be performed to clarify whether the causal relationship between BMI and COVID-19 illness extends to critical respiratory illness. While contributing studies to the Host Genetics Initiative did not provide information on self-reported race or ethnicity, most were presumably European and association analyses were adjusted for ancestry PCs. Well-powered studies in people of non-European ancestral origins are critically needed to as ethnic and racial minorities in the U.S. are disproportionately affected by the pandemic^7,11,27,72-74^. We recognize that the primary social drivers of viral exposure and spread (i.e., crowding within households, wealth and education gaps, working in essential jobs that render social distancing challenging, language barriers, and poor access to healthcare) are likely correlated with, or are themselves, determinants of obesity^75,76^. Future investigations are required to determine whether addressing these upstream social factors mitigates the impact of obesity on COVID-19 outcomes.

## CONCLUSION

Our study provides genetic evidence that support or refute causality for a plethora of cardiometabolic conditions that can inform preventive strategies aimed at modifying risk of COVID-19 illness. Among the 17 cardiometabolic exposures tested, only BMI was found to be a causal risk factor for COVID-19 susceptibility and severity, which is consistent with multiple epidemiologic studies that have reported an association between obesity and COVID-19 illness. We conclude that obesity may have amplified the disease burden of the COVID-19 pandemic either single-handedly or through its metabolic consequences. To the extent that obesity is a modifiable risk factor with a strong environmental component, public health measures that aim to diminish the societal obesity burden could be incorporated into an effective preventive strategy for COVID-19 outcomes. Similarly, preventive measures that increase the risk of obesity (e.g. limitation of access to open spaces for exercise) should be viewed with caution. Future research is required to understand the mechanisms through which obesity increases the risk of COVID-19 outcomes, and whether obesity-related conditions are along the causal pathway. Our study has shown how large-scale genotype-phenotype summary data rapidly assembled during a pandemic and made freely accessible to the research community can accelerate research with immediate and direct application to clinical practice and public health messaging.

## Data Availability

All the results from this study are presented in the manuscript and online supplement. All datasets used to generate results are publicly available.

## DATA ACCESS AND RESPONSIBILITY

Drs. Aaron Leong and Josep M. Mercader had full access to all the data in the study and takes responsibility for the integrity of the data and the accuracy of the data analysis.

## ACKNOWLEDGEMENTS

We thank all the genetics consortia and the COVID-19 Host Genetics Initiative for making summary statistics publicly accessible for this analysis. As supplemental information, we acknowledge the contribution of the studies in the COVID-19 Host Genetics Initiative listed at https://www.covid19hg.org/acknowledgements/ and list all MEGASTROKE authors. The MEGASTROKE project received funding from sources specified at http://www.megastroke.org/acknowledgments.html.

